# Physiological effects of dual target DBS in an individual with Parkinson’s Disease and a sensing-enabled pulse generator

**DOI:** 10.1101/2024.02.23.24303295

**Authors:** Daniel D. Cummins, Stephanie S. Sandoval-Pistorius, Stephanie Cernera, Rodrigo Fernandez-Gajardo, Lauren Hammer, Philip A. Starr

**Author notes:** Shared first authorship (Daniel D. Cummins, Stephanie S. Sandoval-Pistorius). To whom correspondence should be addressed: Daniel D. Cummins, MD Department of Neurosurgery Mount Sinai Health System. **Funding:** This research did not receive any specific grant from funding agencies in the public, commercial, or not-for-profit sectors.

## Abstract

**Introduction:** Deep brain stimulation (DBS) of the subthalamic nucleus (STN) or globus pallidus (GP) is an established therapy for Parkinson’s disease (PD). Novel DBS devices can record local field potential (LFP) physiomarkers from the STN or GP. While beta (13-30 Hz) and gamma (40-90 Hz) STN and GP LFP oscillations correlate with PD motor severity and with therapeutic effects of treatments, STN-GP interactions in electrophysiology in patients with PD are not well characterized.

**Methods:** Simultaneous bilateral STN and GP LFPs were recorded in a patient with PD who received bilateral STN-DBS and GP-DBS. Power spectra in each target and STN-GP coherence were assessed in various ON- and OFF-levodopa and DBS states, both at rest and with voluntary movement.

**Results:** OFF-levodopa and OFF-DBS, beta peaks were present at bilateral STN and GP, coincident with prominent STN-GP beta coherence. Levodopa and dual-target-DBS (simultaneous STN-DBS and GP-DBS) completely suppressed STN-GP coherence. Finely-tuned gamma (FTG) activity at half the stimulation frequency (62.5Hz) was seen in the STN during GP-DBS at rest. To assess the effects of movement on FTG activity, we recorded LFPs during instructed movement. We observed FTG activity in bilateral GP and bilateral STN during contralateral body movements while on GP-DBS and ON-levodopa. No FTG was seen with STN-DBS or dual-target-DBS.

**Conclusion:** Dual-target-DBS and levodopa suppressed STN-GP coherence. FTG throughout the basal ganglia was induced by GP-DBS in the presence of levodopa and movement. This bilateral STN-FTG and GP-FTG corresponded with the least severe bradykinesia state, suggesting a pro-kinetic role for FTG.

## Introduction

Deep brain stimulation (DBS) is an established therapy for Parkinson’s disease (PD). The most common anatomic targets for implantation of leads are the subthalamic nucleus (STN) or globus pallidus (GP). Local field potential (LFP) physiomarkers may be recorded from the leads implanted in STN or GP, and are thought to reflect synaptic currents in neuronal elements close to the recording site. These are often analyzed in the frequency domain, and the spectral power of oscillations in particular frequency bands, including beta (13-30 Hz) and finely-tuned gamma (FTG; 30-90 Hz), correlate with severity of motor signs and with therapeutic effects [1,2].

In complex cases, dual-target DBS (DT-DBS), with leads placed in both the STN and GP, may be used to maximize therapeutic benefit [3]. These cases provide a rare opportunity to study interactions between these basal ganglia nuclei, and effects of stimulation at one target on oscillatory activity at the other target. However, limited work has explored STN and GP LFPs recorded simultaneously. A perioperative study, recording from externalized leads, demonstrated prominent STN-GP beta coherence in the OFF-levodopa state, while STN-GP FTG coherence was more prominent in the ON-levodopa state [4]. However, perioperative recordings are subject to “microlesion” effects of lead insertion. Next-generation bidirectional DBS devices capable of recording chronic LFPs months to years after implantation are enabling further studies of STN, GP, and STN-GP physiology [5–7]. Here, we explore levodopa and DBS effects on STN-GP beta and gamma LFPs using a commercially available bidirectional DBS system.

## Methods

The study subject was a right-handed male with PARK2 juvenile-onset PD (NM_004562.3(PRKN) homozygous c.155 deletion, p.Asn52Metfs*29) who received bilateral STN-DBS (Medtronic 3387 leads) in 2005. Bilateral GP leads (Medtronic B3301542) were added in 2021 to treat worsening ON-OFF fluctuations requiring higher doses of levodopa and increased STN stimulation amplitudes, leading to stimulation-induced dyskinesias. Bilateral GP and bilateral STN leads were connected to separate Percept PC pulse generators (Medtronic B35200) capable of LFP recording. Surgical procedures for DBS implantation have been previously described [8]. ON- and OFF-medication states were characterized using the Movement Disorder Society Unified Parkinson’s Disease Rating Scale motor subscale (MDS-UPDRS Part III). A bradykinesia score was determined by components of the MDS-UPDRS Part III (components 3.4-3.8) including finger tapping, hand opening-closing, hand supination-pronation, toe-tapping, and leg agility. As the subject’s most prominent PD symptom, bradykinesia was the focus of the assessment. They did not experience tremor and symptoms of rigidity and dystonia could not reliably be scored via video. Institutional review board approval was obtained for this work with informed consent obtained from the patient.

The postoperative localization of DBS contacts and volume of tissue activated estimation were performed using the Lead-DBS toolbox and the integrated FieldTrip-SimBio pipeline in MATLAB (**Figure 1A**) [9]. The gradient of the potential distribution was thresholded for magnitudes above 0.2V/mm. Pre- and post-operative MRI scans were co-registered and normalized to the Montreal Neurological Institute (MNI) space (MNI152 NLIN 2009b), and contacts were projected onto the DISTAL Atlas [10].

**Figure 1.**
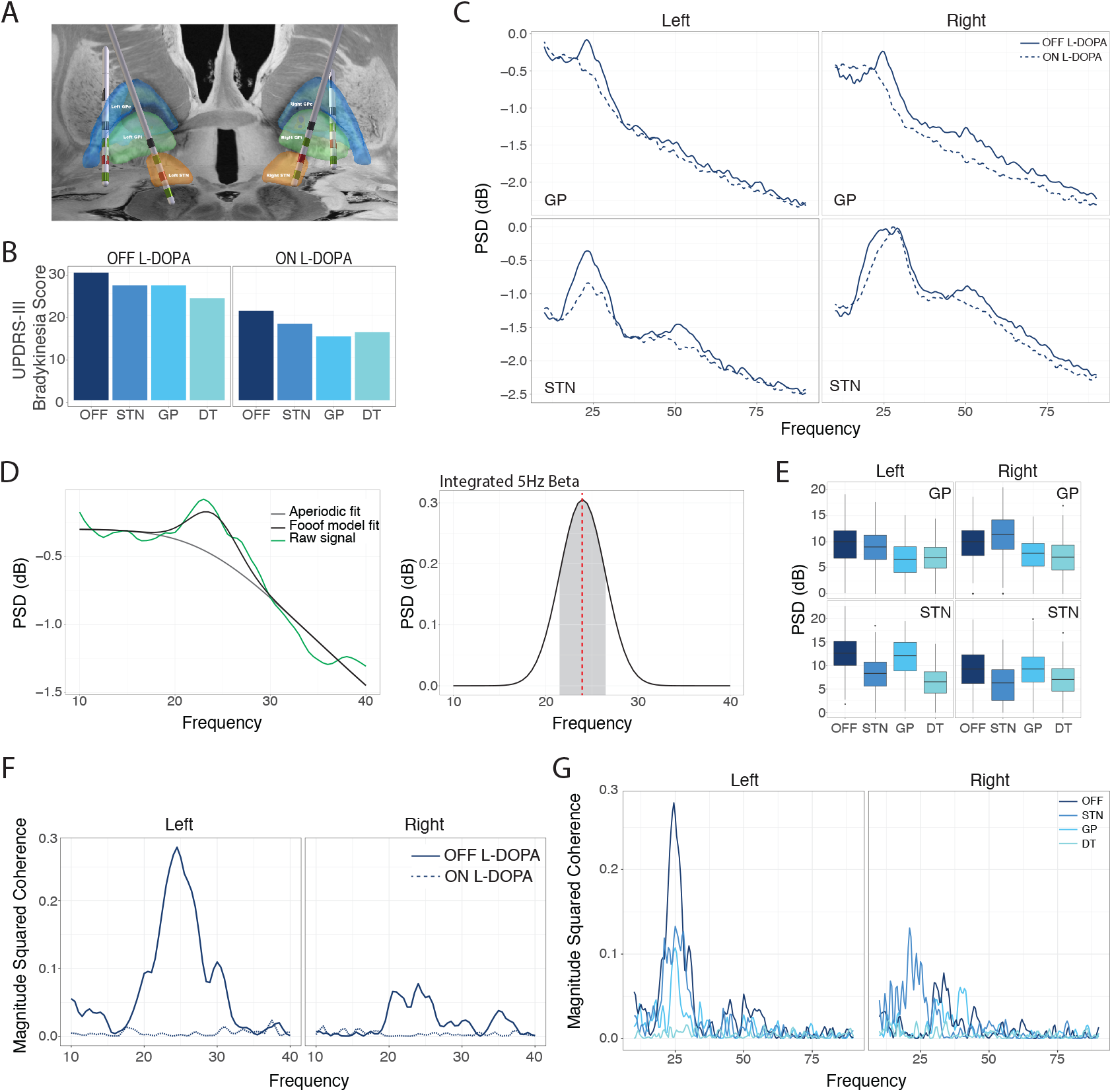
**(A)** LEAD-DBS reconstruction of lead placements in the bilateral STN and GP. Local field potentials were recorded in bipolar configuration from contacts 0-2 at each target. Lead coordinates of the therapeutic contact are given in Montreal Neurological Institute (MNI) space in mm. Compared with the left STN (MNI coordinates X: −12.1, Y: −12.4, Z: −8.0), right STN contacts are shifted more dorsally (MNI coordinates X: 15.7, Y: −12.6, Z: −5.1). Compared with the right GP (MNI coordinates X: 21.9, Y: −4.0, Z: −3.1), left GP leads are shifted more laterally (MNI coordinates X: −22.8, Y: −6.7, Z: −6.1), with greater globus pallidus externus (GPe) lead contact on the left GP compared to the right GP. Red = stimulation contact. Green = sensing contacts. **(B)** Unified Parkinson’s Disease Rating Scale motor subscale-III bradykinesia scores in all L-DOPA and DBS conditions, with the best bradykinesia score in the ON-LDOPA, GP-DBS state and worst in the OFF L-DOPA, OFF-DBS state. **(C)** Power spectral density (PSD) plots around the beta and gamma bands (10Hz-90Hz) from local field potential recordings while ON L-DOPA versus OFF L-DOPA, all OFF-DBS. The patient was instructed to remain at rest during all recordings. Beta activity was reduced at bilateral STN and bilateral GP with administration of L-DOPA **(D)** Method for determining integrated peak beta activity. From the raw PSD plot (green), the full FOOOF (black) and aperiodic component (grey) were calculated. By subtracting the aperiodic component from the full modeled spectrum, the periodic activity was determined (D, right). Integrated peak power was determined in a 5Hz band around the center frequency of peak beta activity. **(E)** Integrated peak beta activity, determined by the method in (D), at bilateral STN and bilateral GP across DBS conditions (OFF L-DOPA). In the OFF-L-DOPA state, DT-DBS and STN-DBS similarly reduced bilateral STN beta peak power compared to OFF-DBS, while GP-DBS minimally reduced STN beta power. Within the GP, DT-DBS and GP-DBS similarly reduced bilateral GP beta power compared to OFF-DBS. STN-DBS had minimal effect on left GP beta, and increased GP beta power in the right hemisphere **(F)** Magnitude-squared coherence between the left STN and left GP demonstrates a coherence peak of 0.28 at 24.5Hz in the OFF-levodopa, OFF-DBS state, not apparent ON-levodopa. A smaller beta peak in coherence OFF-levodopa was also seen between the right STN and right GP. (**G)** In the OFF-L-DOPA state, DT-DBS suppressed bilateral STN-GP beta coherence, with a smaller effect from STN-DBS or GP-DBS alone.

LFPs were recorded simultaneously from both targets ON- and OFF-L-DOPA (6 hours following previous dose) and during single-target STN- or GP-DBS, and DT-DBS. Stimulation frequency was 125Hz and pulse width 60us for all targets. Stimulation amplitude was 2.4mA at bilateral STN and 2.2mA at bilateral GP. Contact 1 (the second most ventral contract) was used for therapeutic stimulation at bilateral STN and bilateral GP. Stimulation parameters at STN and GP were the same for dual-target DBS and single-target DBS. LFPs were recorded from all targets in bipolar configuration (contacts 0-2) flanking the stimulating contact. Sense-friendly STN and GP stimulation parameters optimized for DT-DBS were used for all ON-DBS recordings.

LFPs were sampled at 250Hz and bandpass filtered between 1Hz and 100Hz. LFPs were analyzed in the frequency domain using the Welch method with a Hamming window of one second and 50% overlap (*pwelch.m* and *spectrogram.m* from MATLAB Signal Processing Toolbox). The periodic component of each power spectrum was then isolated around the beta band (10Hz – 40Hz) and gamma band (40Hz – 90Hz) using the previously described fitting one over f (FOOOF) algorithm [11]. Magnitude-squared coherence was determined using Welch’s overlapped averaged periodogram method with a one second window (*mscohere.m* from MATLAB Signal Processing Toolbox).

LFP data from each Percept PC and kinematics data were synchronized by aligning stimulation artifacts detected by Trigno electromyography sensors placed over the DBS extension wires (Delsys, Inc). Except where movement is indicated, all data were recorded with the patient instructed to be at rest. Periods of instructed voluntary movement (administration of the MDS-UPDRS Part III) were confirmed using accelerometry data on bilateral arms and legs that were high-pass filtered at 0.1Hz.

## Results

### Effects of levodopa and DBS on subthalamic and pallidal beta power

Bradykinesia scores demonstrated combined effects of levodopa and DBS, with the least severe bradykinesia score in the ON-LDOPA, GP-DBS state (**Figure 1B**). Previous studies suggest that DT-DBS may be more beneficial to patients than STN- or GP-DBS alone, which may correspond with decreased beta activity [3,7]. To assess the effects of single and DT-DBS on STN and GP beta-band spectral power, we recorded LFPs simultaneously from bilateral STN and GP. Levodopa reduced beta activity at bilateral STN and bilateral GP in the OFF-DBS state (**Figure 1C**). From power spectra at each target, we calculated an area under the beta peak in a 5Hz bandwidth centered on the OFF-L-DOPA/OFF-DBS beta peak (**Figure 1D**). This metric was chosen because it is a long-term trackable parameter in the Percept PC system for potential application in adaptive DBS, in which DBS parameters are modulated based on sensed physiomarkers [12]. In the OFF-L-DOPA state, DT-DBS and STN-DBS similarly reduced bilateral STN beta peak power compared to OFF-DBS, while GP-DBS minimally reduced STN beta-band spectral power (**Figure 1E**). Within the GP, DT-DBS and GP-DBS similarly reduced bilateral GP beta-band spectral power compared to OFF-DBS. STN-DBS had minimal effect on left GP beta, and surprisingly, increased GP beta-band spectral power in the right hemisphere (**Figure 1E**).

### STN-GP beta coherence reduced by dual target DBS

STN-GP beta coherence is increased by dopamine depletion [4]. We analyzed simultaneous STN and GP LFP recordings OFF-L-DOPA, OFF-DBS. We observed prominent STN-GP beta coherence in the left hemisphere, with a 24.5Hz peak **(Figure 1F)**. Right hemisphere STN-GP beta coherence was less prominent, but still present, with a 24.0Hz peak (**Figure 1F**). No STN-GP beta coherence was observed in the ON-L-DOPA, OFF-DBS state. In the OFF-L-DOPA state, DT-DBS also suppressed bilateral STN-GP beta coherence, with a smaller effect from STN-DBS or GP-DBS alone (**Figure 1G**).

### Finely-Tuned Gamma in the basal ganglia is induced with GP-DBS, levodopa, and voluntary movement

FTG oscillatory activity in the cortex and basal ganglia is linked to pro-kinetic states, both with voluntary movement and involuntary dyskinesias [13,14]. We observed entrained FTG activity at half the stimulation frequency (62.5Hz) within the left STN when ON-levodopa, most prominently during GP-DBS (**Figure 2A**). No FTG was seen in the right STN or bilateral GP in any DBS state, during the resting state. To assess the effect of voluntary movement on FTG activity, we recorded LFPs during the administration of the MDS-UPDRS Part III motor exam. ON-L-DOPA and during GP-DBS, we detected prominent movement-related entrained 62.5 Hz FTG activity in the bilateral STN and bilateral GP (**Figure 2B-C**), which was not apparent in states of OFF-DBS, STN-DBS, or DT-DBS (**Figure 2D-E**). STN entrained FTG activity corresponded with voluntary leg movement, while GP entrained FTG activity corresponded with voluntary right arm movement **(Figure 2B-C)**. The only state with bilateral STN- and GP-FTG (ON-L-DOPA, GP-DBS) also corresponded with the lowest MDS-UPDRS Part III bradykinesia score **(Figure 1B)**. In terms of other symptoms, dystonia was present primarily in the OFF-LDOPA state, with mild improvements with single and dual-target DBS. The patient had minimal to no dystonia in the ON-LDOPA state, regardless of DBS. Dyskinesia only occurred in the ON-LDOPA state, which worsened with STN-DBS condition and reduced with GP-DBS. Precise dystonia and dyskinesia scores were not recorded.

**Figure 2.**
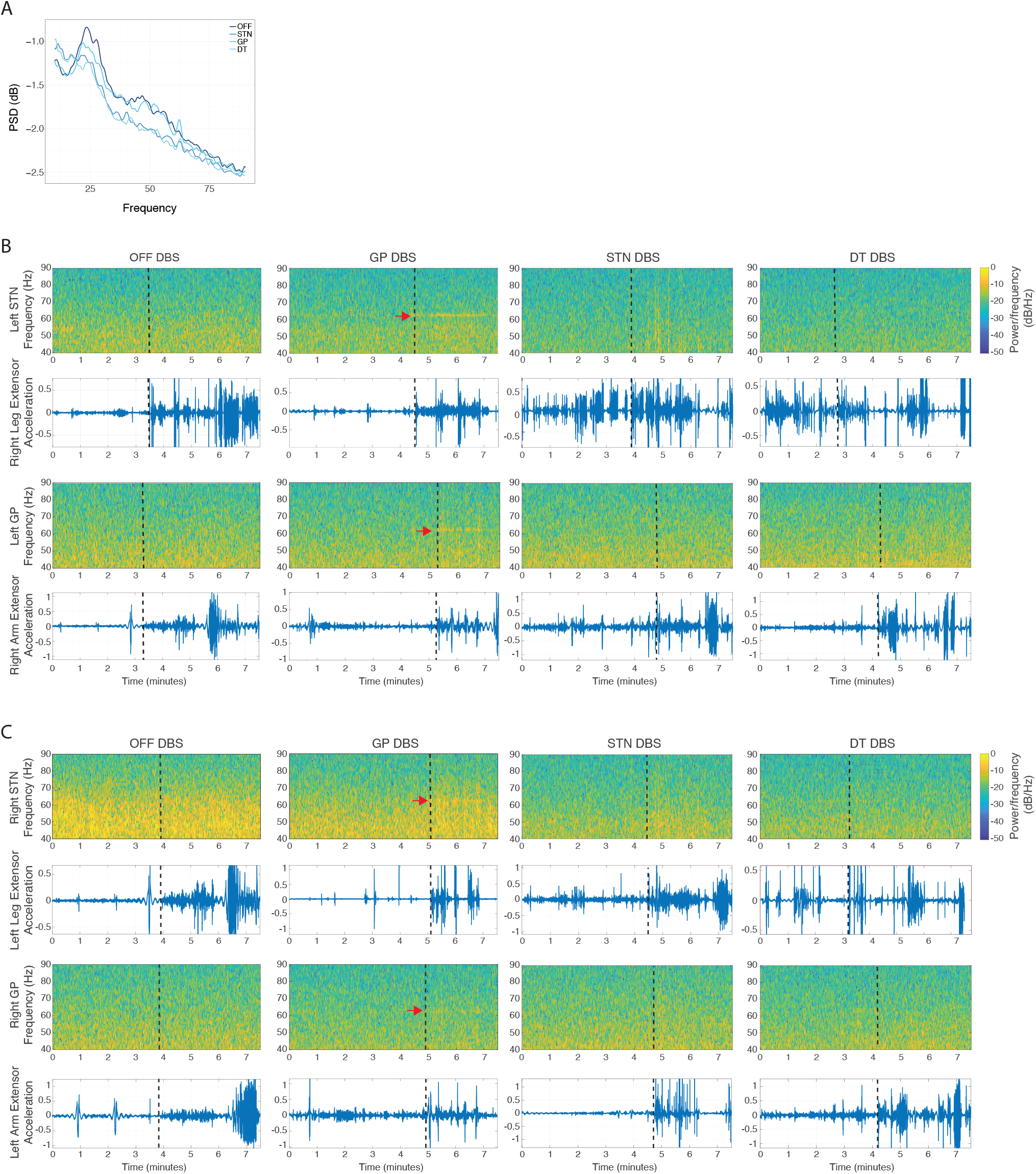
**(A)** Power spectral density plots for left STN in at-rest recordings, ON L-DOPA, in various DBS states. **(B-C)** Spectrograms showing activity over time of recordings in various DBS states while ON-levodopa from the right **(B)** and left **(C)** hemispheres. Corresponding accelerometer data from bilateral upper and lower extremities is also shown to approximate the state of voluntary movement. Finely tuned gamma (FTG) at half the stimulation frequency (62.5Hz) is seen particularly in the STN and GP during GP-DBS. FTG arises with the start of voluntary movement (confirmed with corresponding accelerometry data below). FTG was not seen with STN-DBS or dual-target DBS.

## Discussion

Here, we used a commercially available bidirectional DBS system in a patient with *Parkin*-positive PD, implanted with bilateral pallidal and subthalamic leads, to understand how levodopa and DBS at either target affects oscillatory activity in each target. We demonstrate a combined effect of DT-DBS to reduce STN and GP beta activity. STN-GP coherence in the untreated PD state was shown to be suppressed by both levodopa and DT-DBS. Finally, we show GP-DBS-induced FTG-entrainment in the STN and GP dependent on levodopa and voluntary movement, which matched the state of greatest improvement in bradykinesia.

Both pallidal DBS and subthalamic DBS have been shown to suppress beta activity in the target tissue receiving stimulation, which is postulated to be an important mechanism of the therapeutic effectiveness of DBS [15,16]. Although STN- and GP-DBS have been reported to have similar efficacy on motor signs, the relative efficacy of STN and GP-DBS for beta suppression has not been well evaluated. Our results in one subject suggest that beta suppression in the basal ganglia network occurs at the stimulated target tissue, but not necessarily at functionally related targets (**Figure 1E**). This finding builds upon recent work by *Mitchell et al* showing the greatest effectiveness of beta reduction, at least at STN, with DT-DBS compared to STN- or GP-DBS [7]. There remains limited human literature on STN-GP interactions in PD. Previous studies showing OFF-L-DOPA STN-GP beta coherence were conducted during the peri-operative period, before beginning chronic DBS [14]. We show that STN-GP beta coherence while OFF-L-DOPA persists after years of chronic DBS, corroborating previous findings from acute recordings [14]. While STN-DBS and GP-DBS each reduced beta band spectral power in the stimulated region, DT-DBS was necessary to completely attenuate beta coherence between the two targets (**Figure 1G**) and was associated with improved bradykinesia in the OFF-L-DOPA state compared to single target DBS. Reductions in beta-band spectral power are thought to drive therapy-induced improvements in motor signs. Effects of levodopa or DT-DBS on cross-target interactions, such as STN-GP coherence, may also play a role in motor sign severity. Asymmetry in the magnitude of beta coherence between right and left brain could be related to asymmetries in lead location (**Figure 1A)** or differences in the most affected hemisphere, although he had similar severity of motor signs on each side on the day of testing.

Stimulation-induced entrainment of FTG oscillations by STN-DBS has been reported both in STN and in sensorimotor cortex [18,19]. However, entrainment phenomena between the pallidum and STN have not been studied. We describe the first report of entrainment of both pallidal *and* subthalamic-FTG activity by pallidal DBS, with no such entrainment observed during STN-DBS. Our results build on prior work demonstrating a pro-kinetic role for FTG [2]. Based on canonical basal ganglia networks, these findings also support a possible antidromic mechanism of FTG entrainment, shedding light on the fundamental principles of DBS. A caveat to the presented work is that the patient was not yet fully optimized for GP-DBS at the time of the study and stimulation parameters for STN had to be modified from their most therapeutic setting, for sensing compatibility.

## Data Availability

All data produced in the present study are available upon reasonable request to the authors

